# Communication under sharply degraded auditory input and the “2-sentence” problem

**DOI:** 10.1101/2022.07.22.22277720

**Authors:** Mario A Svirsky, Jonathan D Neukam, Nicole H Capach, Nicole M Amichetti, Annette Lavender, Arthur Wingfield

## Abstract

**Introduction:** Many cochlear implant (CI) users who do quite well in standard clinical tests of speech perception report that a great amount of effort is required when listening in real-world situations. We hypothesize that the combined constraints of the sharply degraded signal provided by a CI and finite cognitive resources may lead to a “tipping point” when listeners are confronted with speech material that is more complex than the single words or single sentences that are used in clinical tests. Beyond this tipping point, communication may become too difficult, even for CI users whose standard speech intelligibility scores are above average.

**Methods:** Here, we investigate whether speech identification performance and processing effort (indexed by pupil dilation measures) are affected when CI users or normal hearing control subjects are asked to repeat two sentences presented sequentially instead of just one sentence.

**Results:** Response accuracy was minimally affected in normal hearing listeners, but CI users showed a wide range of outcomes, from no change to decrements of up to 45 percentage points. The amount of decrement was not predictable from the CI users’ performance in standard clinical tests. Pupillometry measures tracked closely with effort in both the CI group and the normal hearing group, even though the latter had speech perception scores near ceiling levels for all conditions.

**Discussion:** A communicative tipping point may be reached in many (but not all) CI users in response to input that is only minimally more challenging than standard clinical tests; specifically, when two sentences are presented sequentially before requesting a response, instead of presenting just a single sentence at a time. This potential “*2-Sentence Problem*” represents one of the simplest possible scenarios that go beyond presentation of a single word or sentence, and it raises the possibility that even good performers in clinical tests of speech perception may be brought beyond the tipping point by other ecologically relevant manipulations. The present findings also raise the possibility that a clinical version of a 2-sentence test may provide actionable information for counseling and rehabilitating CI users.

## INTRODUCTION

Cochlear implants (CIs) replace the sense of hearing by electronic means and they have had considerable clinical success, not only in children born deaf but also in adults who have lost their hearing after learning language. In adults, “success” is measured clinically using tests of word identification (e.g. Lehiste et al. 1959) or sentence identification (e.g. Spahr et al. 2012), typically in quiet and sometimes in moderate levels of noise. Across countries and languages speech perception tests to evaluate CI users’ performance very rarely (if ever) use linguistic units longer than a single sentence.

Although such tests are important, most human communication takes place in discourse segments longer than one sentence. This raises concerns about the ecological validity of the use of single-word and sentence identification tests. These concerns are particularly acute when there is reason to believe that performance in these clinical tests might not be representative of a listener’s real-world performance. In fact, some CI users complain about the great effort it takes them to communicate in everyday discourse. The social media comments from three CI users shown in Table 1 illustrate the real-world difficulties many CI users experience, and the “listening fatigue” that can lead to withdrawal from social interaction. Indeed, clinicians can often be puzzled by such comments from patients who seem to do well in clinical tests but report major difficulties in everyday interactions.

**Table 1.**
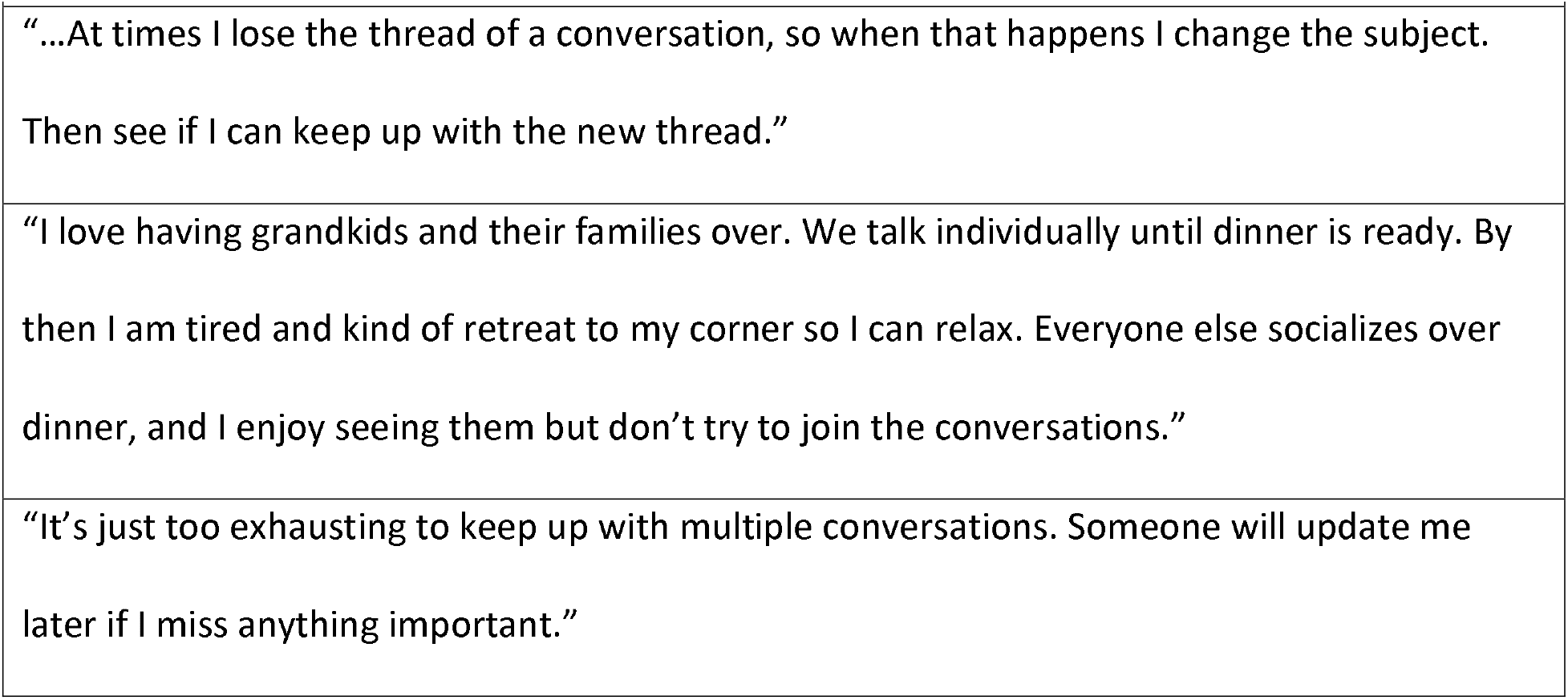
Social Media Comments from CI Users.

We hypothesize that the combined constraints of a sharply degraded auditory input and finite cognitive resources (e.g. Kahneman 1973), bring many (but not all) CI users beyond a “tipping point,” where real-world oral communication becomes too difficult. Importantly, we suggest that this may happen even in patients with above-average scores on traditional clinical speech perception tests. This postulate is aligned with the *effortfulness hypothesis* first proposed by Rabbitt (1991) and later developed by Pichora-Fuller and colleagues’ (2016) Framework for Effortful Listening (FUEL).

As applied to the case of CI users, the concern is that the effort required for successful perception of degraded speech may draw resources that would otherwise be available for post-perceptual operations such as encoding what has been heard in memory. Crucially, this detrimental effect of perceptual effort on speech recall would occur even when the words themselves could be correctly identified (Cousins et al. 2014; McCoy et al. 2005). This is an effect that might be more apparent when CI users are confronted with continuous discourse uttered at normal speech rates than when they have to process the single words or sentences that are used in clinical tests.

In the present study we explore the extent to which the communicative tipping point may be reached by input that is minimally more challenging than standard clinical tests; specifically, when two sentences are presented sequentially before requesting a response, instead of presenting just a single sentence at a time. This potential *“2-sentence problem*” represents one of the simplest possible scenarios that go beyond presentation of a single word or sentence.

This study takes advantage of the fact that CI users represent a uniquely important platform to explore the interaction between cognitive and sensory limitations. This is so for several reasons. First, these listeners experience a very significant degree of auditory input degradation, along with long term exposure to this sharply degraded sensory input. Thus, the present data will help answer the general question of how much signal degradation the human brain may or may not be able to handle even after tens of thousands of hours of listening experience that may help mitigate such signal degradation. Additionally, CI users are an important clinical population and these results may help guide their treatment and rehabilitation.

We report the results of two experiments. The first experiment was an exploratory behavioral study to determine the existence and extent of the 2-sentence problem in CI users. It included three conditions: repeating one sentence (*1-sentence* condition); repeating a sequence of two sentences (*2-sentences-repeat both* condition); and presentation of two sentences followed by a prompt to repeat either the first or the second sentence without advance knowledge of which sentence would be selected (2*-sentences+post prompt*). Because participants do not know in advance which sentence they will be prompted to repeat in the 2*-sentences+post prompt* condition, they have to process both sentences in this condition, even though only one will be repeated.

Once the first experiment established that a 2-sentence problem does indeed exist, a second experiment was conducted that included several modifications. First, participants received the original three conditions plus an additional one: presenting two sentences with listeners <u>knowing in advance</u> that they only had to repeat either the first sentence or the second sentence (*2-sentences+pre-<u>prompt</u>*). The difference between the *2-sentences+pre-prompt* condition and the *2-sentences+*<u>*post prompt*</u> condition is that, in the former, listeners hear the prompt prior to presentation of the sentences (thus “pre-prompt”) and in the latter they hear the prompt some time after the two sentences have been presented (thus “post prompt”). Despite the similarity, the task required from listeners has different implications. In the *2-sentences+pre-prompt* condition they know a priori that they need to focus on a single target sentence and they may disregard the other one whereas in the *2-sentences+post prompt* condition both sentences have to be processed and kept in memory until the “post prompt” is presented. The *2-sentences+pre-prompt* condition was added to explore the extent to which different types and levels of task difficulty influence speech recognition accuracy, listening effort, and their interaction.

Because this study aimed to explore the interaction of degraded sensory input and the investment of cognitive resources in the task at hand, the second experiment also included pupillometry as a measure of processing effort. Pupil dilation responses were obtained starting at the beginning of each stimulus presentation, through the course of a brief retention interval, and continuing until subject response. Along with its response to changes in ambient light (Wang et al. 2006), and response to emotional arousal (Kim et al. 2000; Kinner et al. 2017). Pupil dilation has been increasingly used as an index of cognitive effort in response to a wide range of perceptual and cognitive challenges. These have included a transient increase in pupil dilation accompanying tasks including perception of speech degraded by noise (Zekveld et al. 2010), or mild-to-moderate hearing loss (Ayasse et al. 2018), perception of syntactically simple vs. complex sentences (Piquado et al. 2010), low-context sentences vs. high-context sentences (Winn 2016), perception of spectrally degraded speech (Wagner et al. 2016; Winn et al. 2015), and perception of two simultaneous sentences (Koelewijn et al. 2015).

It is important to note that, in addition to using pupil dilation as an index of listening effort while a sentence is being heard, useful data can often also be obtained by examining the pupillary response that continues for a period after a sentence is heard (Winn and Moore 2018). For this reason, in Experiment 2 a continuous measure of pupil dilation was obtained starting at the beginning of each stimulus presentation, through the course of a brief retention interval, and continuing until the participant responds. Examination of the dynamic changes in the pupillary response during the course of a behavioral task may thus help illuminate the underlying cognitive processes as they evolve.

Lastly, the second experiment included a comparison group of normal hearing listeners.

In summary, the experiments reported here explore the extent to which a “2-sentence problem” exists for cochlear implant users, whether this behavioral challenge is reflected in pupillary responses, and what inferences we might be able to draw from the combined behavioral and physiological data set.

## EXPERIMENT 1

### Participants

Participants were 14 adult CI users with ages ranging from 25 to 78 years (*M* = 54, *SD*=16.9). Of the 14, 8 were unilateral CI users (and 4 of these were classified as single-sided deaf because they had near normal hearing in the non-implanted ear), 5 were bilateral CI users, and 1 was a “bimodal” CI user, that is, someone who used a conventional hearing aid in the contralateral ear (See Table 2 for a summary of participant demographics).

**Table 2:** Participant demographics for Experiment 1.

| SUBJECT | AGE | GENDER | MODALITY | DEVICE | PROCESSOR | YEARS POST INITIAL STIM | ETIOLOGY |
| --- | --- | --- | --- | --- | --- | --- | --- |
| CI1 | 20s | F | bilateral | CI24RE/CI24R | N5/N5 | 14.8/11.0 | Congenital |
| CI2 | 20s | M | unilateral | CI22M/- | N6/- | 27.0/- | Congenital |
| CI3 | 30s | M | bilateral | HiFocus 1j/HiFocus 1j | Naida/Naida | 13.1/6.7 | Unknown |
| CI4 | 40s | M | unilateral | -/CI24M | -/N5 | -/21.9 | Meningitis |
| CI5 | 40s | F | SSD | CI532/- | N6/- | 0.75/- | SNHL |
| CI6 | 50s | F | unilateral | CI24RE (CA)/- | N6/- | 10.7/- | Cholesteatoma |
| CI7 | 50s | F | SSD | CI532/- | N7/- | 0.09/- | SNHL |
| CI8 | 50s | F | SSD | Flex28/- | Sonnet/- | 0.16/- | SNHL |
| CI9 | 60s | F | bilateral | CI24RE/CI512 | N6/N6 | 13.0/8.9 | Unknown |
| CI10 | 70s | F | SSD | -/Flex28 | -/Sonnet | -/0.17 | NF2 |
| CI11 | 70s | F | bilateral | HiFocus 1j/HiFocus 1j | Naida/Naida | 21.2/10.2 | Unknown Progressive |
| CI12 | 70s | M | bilateral | CI24RE/CI24RE | N7/N7 | 5.7/2.2 | Congenital |
| CI13 | 70s | M | bimodal | -/Flex28 | -/Sonnet | -/1.4 | Unknown Progressive |
| CI14 | 70s | F | unilateral | -/HiFocus 1j | -/Naida | -/2.9 | Familial |

All participants (in both experiments) were recruited and tested in accordance with IRB approval # 18-01806.

### Stimuli and Procedures

The stimuli consisted of 100, 4-to 12-word sentences taken from lists 18 to 23 of the Minimum Speech Test Battery (MTSB) from AzBio speech corpus (Auditory Potential LLC, Goodyear, Arizona USA). These lists were selected because they are commonly not used in a clinical testing (Spahr et al. 2012), and our participants would be unlikely to be familiar with them. Each of the sentences was recorded onto computer sound files (16 bit) at a sampling rate of 44.1 kHz.

Stimuli were prepared for three conditions that would be tested in the experiment:

*1-sentence*. Participants received 20 sentences, one at a time, with instructions to repeat the sentence aloud as accurately as possible.

*2-sentences-repeat both*. Participants received 10 sets of two sentences, with a 500 ms silent pause between the two sentences in a set. Participants were told (*a priori*) that, after both sentences of a pair had been presented, they were to repeat aloud both sentences.

*2-sentences+post prompt*. Participants received 10 sentence pairs (just like in the *2-sentences-repeat both* condition), again with the two sentences in a pair separated by a 500ms silent period. In this case, however, they were told that, after both sentences of a pair had been presented, they would receive a prompt instructing them to repeat either the first or the second sentence. The name of this condition includes the phrase “post prompt” to indicate that listeners heard the prompt *after* having heard the two sentences. The first and second sentence repeat prompts were randomly inter-mixed. Crucially, because participants would not know in advance which sentence they would be asked to repeat, they would have to process and retain in memory both sentences.

Participants were tested in a sound attenuating booth. To isolate the implanted ear and avoid ceiling effects, the four subjects who had near normal hearing in one ear received stimuli via direct audio input to their speech processor. For all other subjects, stimuli were presented through a sound field loudspeaker positioned at zero azimuth, 1m away from the listener. The stimuli were presented at a mean sound level of 65 dB (C scale), which was reported to be a comfortable listening level by all of the participants. Testing was conducted with participants’ everyday sound processor settings. The conditions (*1-sentence, 2-sentences-repeat both*, and *2-sentences+post prompt*) were blocked in presentation, with the order of blocks counterbalanced across participants to minimize practice or fatigue effects. Instructions for each condition were given prior to the condition block (e.g., for the *2-sentences-repeat both* condition the instructions would be “You will hear two sentences. Please repeat back both sentences”).

### Data analysis

Speech scores were analyzed with a one-way, repeated measures ANOVA as a function of testing condition with three levels (*1-sentence, 2-sentences-repeat both, and 2-sentences+post prompt*). Tukey’s multiple comparisons test was used for post-hoc comparisons among all pairs of conditions. Reported *p*-values were multiplicity-adjusted. Sphericity was not assumed by default. Instead, the Geisser-Greenhouse correction was used which is why some of the results that follow may show non-integer degrees-of-freedom values.

### Results

Figure 1 shows the mean percentage of words reported correctly for each condition of Experiment 1. As suggested by the pattern in Figure 1, the ANOVA confirmed a significant effect of conditions on speech accuracy (F [1.411, 18.34] = 13.32, *p* < 0.001, η_p_^2^ = 0.51).

**Figure 1.**
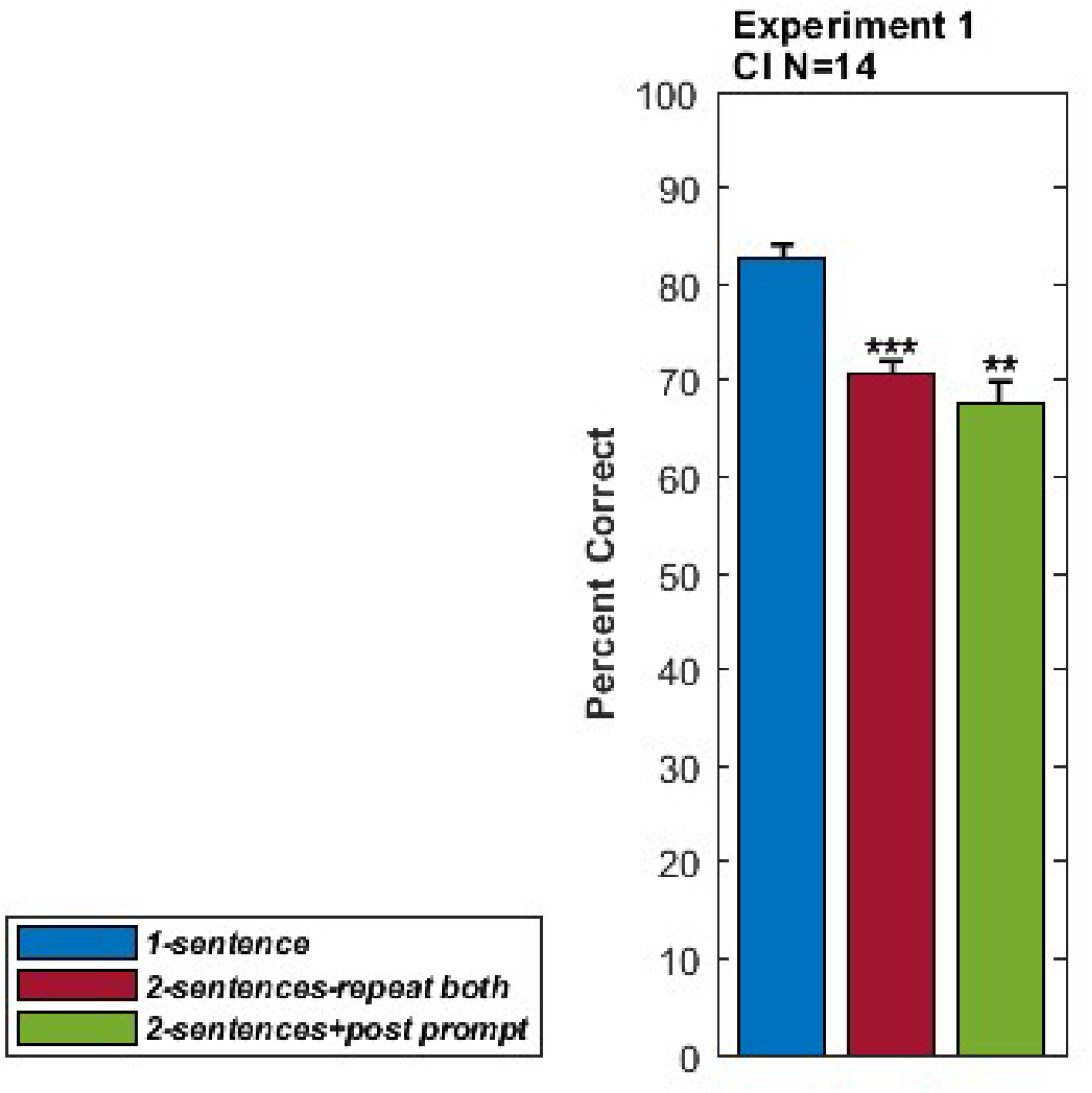
Percent correct responses for CI users for each condition in Experiment 1. Error bars show within-subject standard errors (Loftus et al. 1994). *Asterisks indicate conditions that resulted in significantly worse performance relative to the 1-sentence condition (**=p<0*.*01, ***=p<0*.*001)*.

Table 3 shows the individual participants’ accuracy scores for each of the three test conditions. The final column shows the “2-sentence decrement” in the form of the change in percentage points from the one-sentence condition (the easiest condition) to the *2-sentences+post prompt* condition (the hardest condition). Pairwise comparisons after ANOVA confirmed that having to process and repeat two sentences (the *2-sentences-repeat both* condition) produced a significant decrement of 12 percentage points relative to the *1-sentence* condition (*p* < 0.001). There was also a significant decrement in the *2-sentences+post prompt* condition (14.9 percentage points) relative to the *1-sentence* condition (*p* < 0.01). Differences among conditions remained significant (p<0.01 and p<0.001, respectively) even after removing subject CI9, the one with the most pronounced 2-sentence decrement, from the analysis. In other words, the 2-sentence decrement in this data set was a robust effect that was not driven by a single outlier.

**Table 3.** Individual Differences in 2-sentence decrement, Experiment 1. Two-sentence decrements shown are based on the difference between the *1-sentence* score and the *2-sentences+post prompt* score. Cells in this column are color coded to indicate the magnitude of 2-sentence decrement (green: no decrement; orange-red: large decrement).

| Subjects | 1 Sentence | 2 Sentences | 2 Sent + Post-Prompt | 2-sentence drop |
| --- | --- | --- | --- | --- |
| CI8 | 50.3% | 42.5% | 55.2% | -4.9% |
| CI3 | 92.5% | 82.5% | 95.0% | -2.5% |
| CI4 | 80.3% | 74.8% | 78.1% | 2.2% |
| CI13 | 96.9% | 84.2% | 92.5% | 4.3% |
| CI7 | 79.6% | 69.2% | 69.7% | 9.8% |
| CI10 | 31.5% | 35.8% | 20.8% | 10.7% |
| CI14 | 90.6% | 69.9% | 79.4% | 11.2% |
| CI5 | 100.0% | 83.2% | 84.4% | 15.6% |
| CI11 | 98.7% | 88.4% | 80.6% | 18.1% |
| CI2 | 86.2% | 71.2% | 67.6% | 18.6% |
| CI6 | 77.4% | 62.3% | 56.1% | 21.3% |
| CI1 | 89.7% | 71.5% | 63.1% | 26.6% |
| CI12 | 86.9% | 81.1% | 54.2% | 32.7% |
| CI9 | 94.9% | 73.0% | 50.0% | 44.9% |
| AVERAGE | 82.5% | 70.7% | 67.6% | 14.9% |

### Discussion

There are three major steps in the ability to repeat a spoken sentence. The first is perceptual encoding of the acoustic stimulus, the second is temporarily holding the one (or two) sentences within a limited capacity working memory system (see review in Wingfield 2016), and the third is the retrieval and output of the sentence (or two sentences). The decrement seen in the *2-sentences-repeat both* condition relative to the *1-sentence condition* can be attributed to the increased perceptual and cognitive load represented by all three of these operations. The *2-sentences+post prompt* condition shares the first two elements (encoding and storage) with the *2-sentences-repeat both* condition. This is so because in the *2-sentences+post prompt* condition, not knowing which sentence will be prompted requires that both sentences be perceptually processed and temporarily stored in memory.

The results of this experiment confirm that CI users’ level of success in repeating a single sentence, as commonly tested in the clinic, may underestimate that individual’s difficulty when just one extra sentence is added to the sentence repetition task. Although the effect of this additional load was seen to vary widely from participant-to-participant, in one extreme case adding just a second sentence produced a 2-sentence decrement approaching 45 percentage points that would not have been anticipated by using lists of single AzBio sentences as test material.

## EXPERIMENT 2

After having established in Experiment 1 that CI users experienced a decrease in speech perception accuracy when having to process two sentences at a time, a second experiment was conducted to replicate and extend these initial results. This extension involved an additional participant group composed of normal hearing listeners, one additional experimental condition, and the addition of pupillometry measures obtained during and following stimulus presentation.

A question not addressed in Experiment 1 was whether, even when participants are successful in two-sentence processing, the need to process a second sentence may have had a significant impact on processing effort. To address this question, we added pupillometry to the behavioral task as a measure of processing effort (van der Wel et al. 2018; Zekveld et al. 2018). Although the concept of effort is itself complex (e.g., McGarrigle et al. 2014), for our present purpose we follow Pichora-Fuller and colleagues’ (2016) in defining *effort* as the intentional allocation of attentional or cognitive resources to the accomplishment of a particular task or goal. (See Herrman et al. 2020 for a model of listening engagement; See Peelle 2018 for a detailed discussion of effort and neurocognitive resources.)

### Participants

Twenty-one CI users ranging in age from 19 to 82 (M = 53.8 years, SD = 21), were tested in Experiment 2. Five of the participants were unilateral CI users and 16 were bilateral. Two of the participants with unilateral fittings had residual hearing in the non-implanted ear, with PTAs of 90 and 88dB HL and clinical CNC scores of 4% and 24%, respectively, in those non-implanted ears. All of the CI participants were screened to ensure that their most recent CNC word identification score (Lehiste and Peterson 1959) obtained during the course of their clinical treatment (clinical CNC scores) was at least 60% in the best aided condition. This was done to minimize the likelihood of floor effects in the most difficult conditions.

The CI users’ demographic information is given in Table 4. It should be noted that the CNC30 scores shown in Table 4 are a different recording of monosyllabic words than the CNC recorded words that are used clinically. CNC30 scores are typically 22 percentage points lower than those obtained with the closely related CNC word identification test that is used clinically (Skinner et al. 2006). (Participant CI12 had also taken part in Experiment 1, whereas all other participants had not).

**Table 4.**
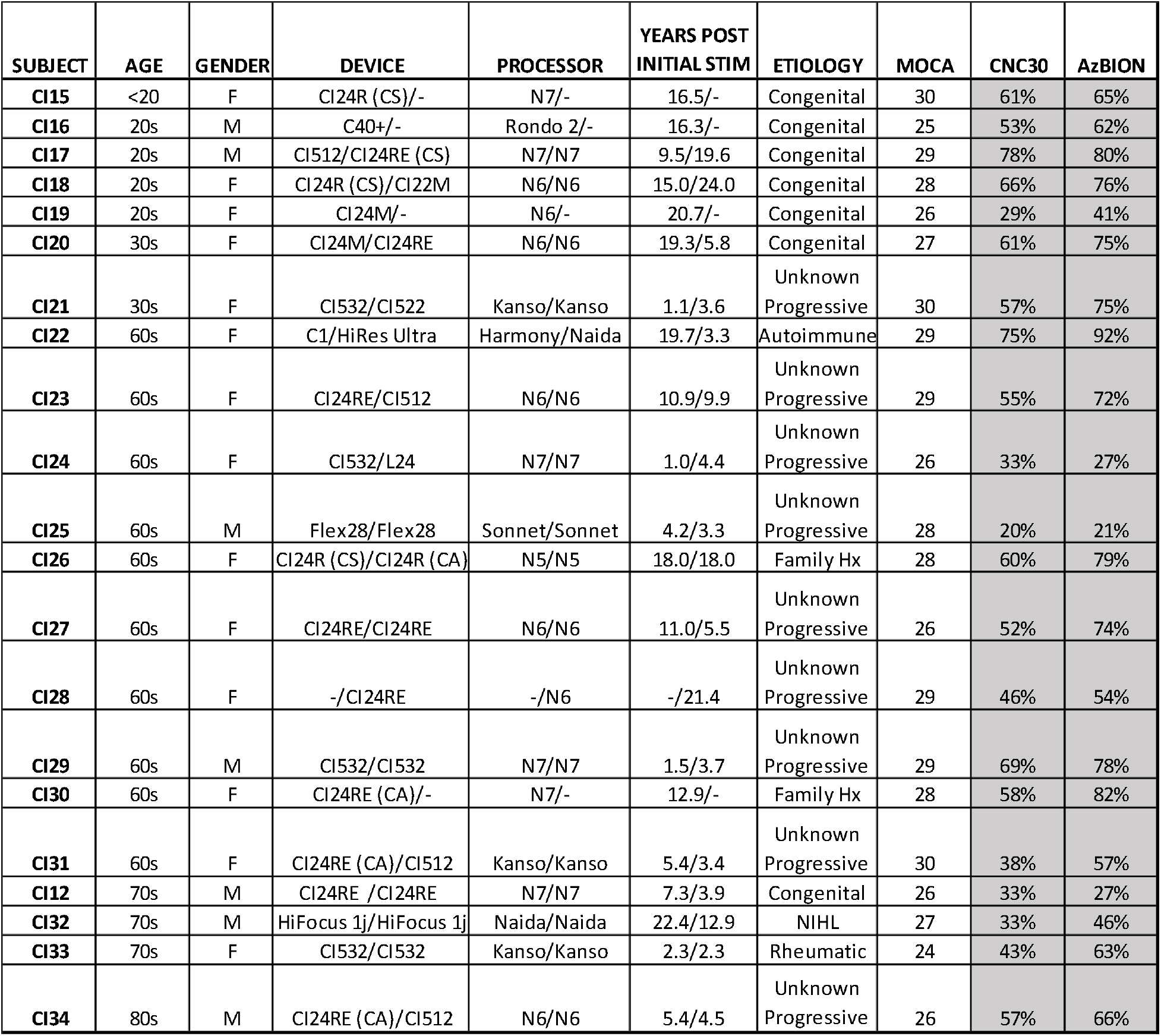
Participant demographics for Experiment 2.

In addition to the CI users, 12 normal hearing adults ranging in age from 21 to 75 (*M* = 43.3 years, *SD* =17.6) were also tested. Hearing was screened at 25dB HL for frequencies of 250, 500, 1k, 2k, and 4k Hz for participants less than 40 years of age (N=6). For participants older than 40 years of age, thresholds were measured from 250 to 8k Hz. The high-frequency pure tone average (1k, 2k, and 4k Hz) for these participants was 17.7dB HL. No participant had a loss greater than 40dB HL at 4k Hz.

### Baseline battery

Several tests were administered to explore whether they might help explain the results. The CI users received two speech perception tests, including two 50-word lists of CNC30 words (Skinner et al. 2006) and two lists of AzBio sentences in noise (Spahr et al. 2012), as well as the Montreal Cognitive Assessment (Nasreddine et al. 2005). Scores for these tests are also shown in Table 4. Three non-speech auditory tests were also administered to the CI users to assess the number of spectral ripples per octave that a listener can detect (SMRT; Aronoff et al. 2013), amplitude modulation detection thresholds (MDT; Landsberger et al. 2021), and temporal gap detection thresholds (Sagi et al. 2009).

Both the CI users and the normal hearing participants received a 20-item version of the Shipley vocabulary test (Shipley 1940).This is a written multiple-choice test in which the participant is required to indicate which of six listed words mean the same or nearly the same as a given target word. The Trails B test (Reitan et al. 1995) was used to assess executive function and inhibition. In this test, participants are asked to create numeric, alphabetical, and alpha-numeric sequences with mouse clicks.

### Stimuli and Procedures

Recorded AzBio sentences as described in Experiment 1 served as stimuli. All three conditions in Experiment 1 were again used in Experiment 2 (*1-sentence, 2-sentences-repeat both*, and *2-sentences+post prompt*. An additional condition was added in which participants received a set of two sentences and were instructed beforehand to repeat either the first or the second one (*2-sentences+pre-prompt condition*). The difference with the *2-sentences+post prompt* condition is that participants were told in advance which of the two sentences they were to repeat. It can be seen in this condition that the memory load would presumably be the same as in the 1-sentence condition. Relative to the 1-sentence condition, however, the act of disregarding the not-called-for sentence might require increased effort for a successful response.

Experiment 2 was self-paced, with the participant using a mouse-click to initiate each subsequent trial. Each trial began with visually presented instructions for that condition followed by another mouse-click, a 500 ms silent period to serve as a pre-sentence pupillometry baseline, and then the stimulus sentence(s). Following the sentence(s) there was a three second silent pause followed by an audible tone as a signal to give the response. In the case of the *2-sentences+post prompt* condition the three second pause included a verbal prompt (starting at 1.8 seconds approximately) indicating whether to repeat back the first or second sentence. As in Experiment 1, the conditions were blocked in presentation, with the order of condition blocks varied between participants using a Latin square design to minimize practice or fatigue effects.

### Pupillometry Data Collection

Pupil sizes were continuously recorded throughout each trial via a desktop-mounted Eyelink 1000 Plus eye-tracker (SR Research, Ontario, Canada), using a standard 9-point calibration procedure. Pupillary data were processed in Matlab (MathWorks, Natick, MA, USA).

An adjustable chin rest was used to minimize head movements and to maintain a distance of 60 cm from the EyeLink camera. To further facilitate reliable pupil size measurements participants were instructed to keep their eyes on a centrally located fixation cross that was continuously displayed on a computer screen placed above the EyeLink camera. The computer screen was filled with a medium gray color to avoid the ceiling or floor effects in the pupil size at baseline (Winn, Wendt, et al. 2018). Ambient light was kept constant throughout the experiment.

Pupil sizes were baseline corrected to account for non-task changes in the base pupil size as can occur across trials (Ayasse et al. 2020). Following the procedures outlined by Reilley and colleagues (2019) this was accomplished by subtracting the mean pupil size measured over the 500 ms baseline silent period preceding each sentence, from the task-related pupil size measures. Eye blinks were detected and removed using the algorithm described by Hershman et al. (2018) and filled by means of linear interpolation.

An additional adjustment was made to account for the tendency for the older adults’ base pupil size and dynamic range to be smaller than that for younger adults (senile miosis; Bitsios et al. 1996; Guillon M 2016). Without such an adjustment, one might underestimate the level of effort allocated to a task by older relative to younger participants. For this purpose we employed light-range normalization, which involved representing pupil sizes as a percentage ratio of the individual’s minimum pupil constriction and maximum pupil dilation. Following initial calibration, a 45-s dark screen (0.4 cd/m^2^) was presented followed by a 45-s light screen (199.8 cd/m^2^) to estimate a participant’s maximum and minimum pupil sizes. Each participant’s dynamic range was calculated as: *D*_max_*-D*_min_, where *D*_max_ was the participant’s average maximum dilation over the last 30 seconds of viewing the dark screen and *D*_min_ was the participant’s average minimum dilation over the last 30 seconds of viewing of the light screen (e.g. Ayasse et al. 2016; Ayasse and Wingfield 2018; Winn, Wendt, et al. 2018). Seven participants received calibration screens both before and after the experiment, and in their case the average of the before- and after-experiment calibrations were used for dynamic range determination.

In Experiment 2 one CI participant did not do the behavioral task with pupillometry, and one CI participant’s pupil data were not used due to excessive blinking throughout the task. A trial rejection threshold was set to 40% such that any trial that consisted of more than 40% of blink removal was discarded and not included in analysis. On average, trial rejection from the 19 CI participants was 17.7%. Two normal hearing participants’ data were lost, one due to equipment error and the other due to a corrupted data file. On average, trial rejection from the remaining 10 normal hearing participants was 2.6%.

### Data Analysis

#### Sentence repetition-Experiment 2

Speech scores from Experiment 2 were analyzed with a two-way, mixed-effects ANOVA, where the within-subjects factor was the testing condition (now with four levels, namely *1-sentence, 2-sentences-repeat both, 2-sentences+pre-prompt*, and *2-sentences+post prompt*). The between-subjects factor was subject group (with two levels: CI users versus normal hearing adults). For this analysis as well as for all the other mixed-effects ANOVAs we describe below, Dunnett’s multiple comparisons test was run, with individual variances computed for each comparison, to determine whether speech scores in any condition were lower than those in the *1-sentence* condition (which was used as a reference). In a second post-hoc analysis, Sidak’s multiple comparisons test was used to examine differences among all pairs of conditions, not just against the *1-sentence* condition. As in Experiment 1, sphericity was not assumed by default. The Geisser-Greenhouse correction was used instead.

A second analysis was done for the speech perception data from Experiment 2, including only the CI users. For this analysis, CI users were classified into three subgroups according to their “2-sentence decrement” (defined as the difference in speech perception scores between the *1-sentence* condition and the *2-sentences+post prompt* condition). This is a reasonable but arbitrary definition, in the sense that the 2-sentence decrement could have been defined instead, for example, as the difference between the *1-sentence* condition and the *2-sentences-repeat both* condition. However, this alternative definition would not have influenced the results of any of the analyses that follow.

As observed in Experiment 1, there was a wide range of individual differences in 2-sentence decrement. In the present case we defined three subgroups: a *no decrement* subgroup (N=8) that included CI users whose 2-sentence decrement was under ten percentage points; a *moderate decrement* subgroup (N=5) that included participants whose 2-sentence decrement was between 10 and 20 percentage points; and a *large decrement* subgroup (N=8) that included participants whose 2-sentence decrement exceeded 20 percentage points. These data were analyzed with a two-way, mixed-effects ANOVA, where the within-participants factor was the testing condition (with the same five levels as described above) and the between-participants factor composed of the three decrement levels listed above: no decrement, moderate decrement, and large decrement groups.

Correlations between the 2-sentence decrement and several demographic variables as well as all variables in the baseline battery were calculated (Pearson correlations for continuous variables and point biserial correlations for dichotomous variables such as gender).

#### Pupillometry

The raw data for statistical analyses of the pupillometry data were the peak pupil dilations that occurred within a time window that included the stimulus presentation and the following 3000 ms, obtained from the average time series curve for each subject and condition. These data were examined with the same mixed-effects 2-way ANOVA described above for the speech perception data in Experiment 2. The data set included nineteen CI users and ten listeners with normal hearing, all of whom were evaluated in the five testing conditions listed above.

### Results

#### Sentence repetition accuracy

The speech results are summarized in Figure 2, which shows the mean percentage of words correctly repeated for each condition, with the left panel showing these data for the CI users and the right panel showing the data for the normal hearing participants. The differences found in Experiment 1 between *1-sentence* scores and *2-sentences-repeat both* or *2-sentences+post prompt* scores were clearly replicated in this second, larger sample.

**Figure 2.**
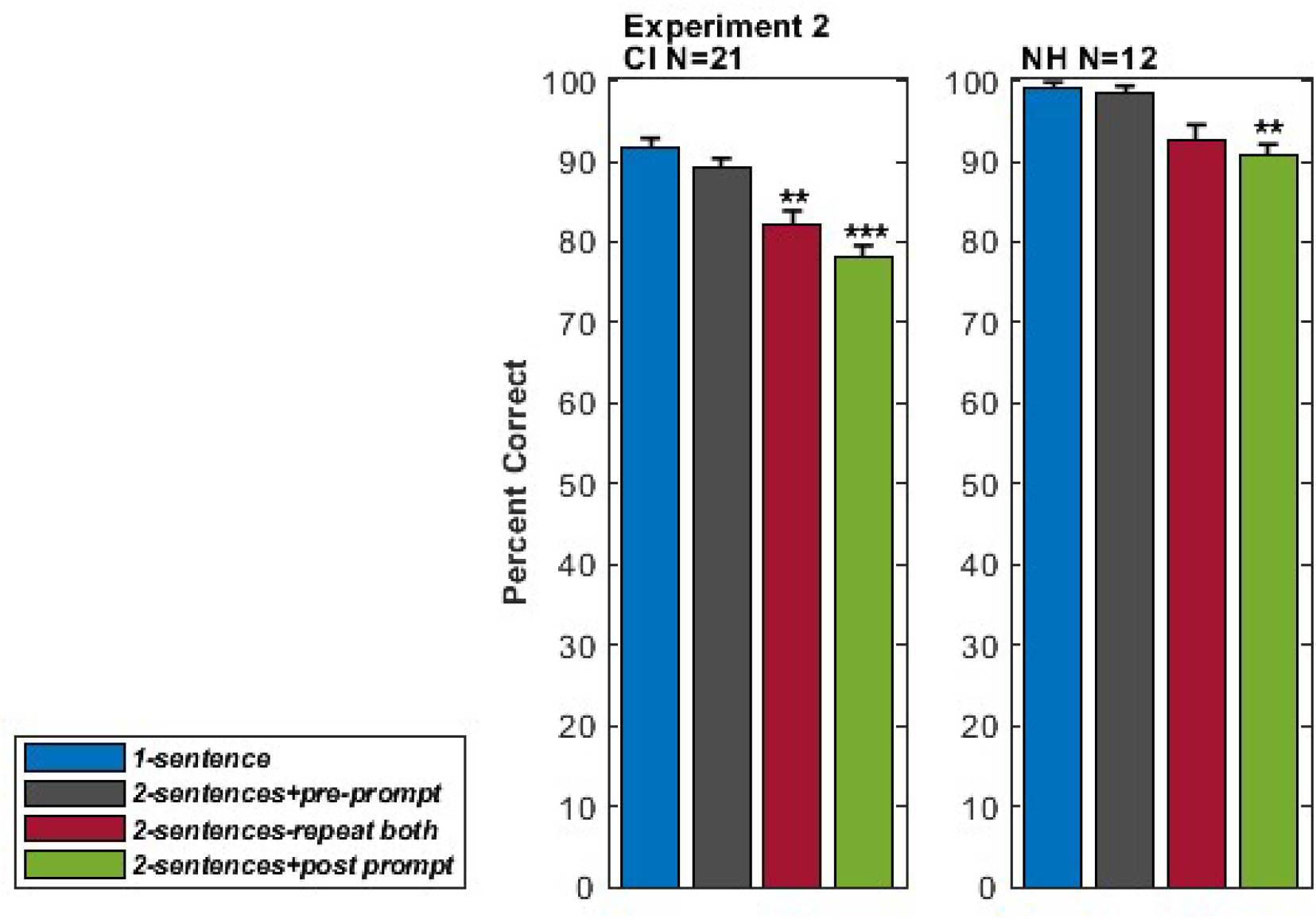
Percent correct responses for CI users (left panel) and normal hearing listeners (right panel). Error bars show within-subject standard errors (Loftus and Masson 1994). Asterisks indicate conditions that resulted in significantly worse performance than the *1-sentence* condition (**=p<0.01, ***=p<0.001).

The appearance of overall better performance by the normal hearing group relative to the CI users was confirmed by a significant main effect of participant group (F [1, 31] = 7.271, p<0.01, η_p_^2^ = 0.44). There was also a significant main effect of condition (F (2.378, 73.71) = 20.15, p<0.001, η_p_^2^ = 0.39) but no significant group-condition interaction, consistent with the observed similarity in the pattern of condition effects for both the CI users and normal hearing participants.

As also found in Experiment 1, scores were highest in the *1-sentence* condition. For the CI group *1-sentence* scores were on average 13.5 percentage points higher than *2-sentences+post prompt* scores (p<0.001) and 9.4 percentage points higher than the *2-sentences-repeat both* scores (p<0.01). For the normal hearing group, *1-sentence* scores were on average 8.4 percentage points higher than *2-sentences+post prompt* scores (p<0.01) and 6.6 percentage points higher than the *2-sentences-repeat both* scores (ns). There were no significant differences between *1-sentence* scores and *2-sentences+pre-prompt* scores, for either the CI group or the normal hearing group. Not surprisingly, speech scores were near ceiling for the normal hearing group, and 9.9 percentage points higher than for the CI group (p<0.05).

In a second post-hoc analysis we compared all conditions against each other instead of only using the*1-sentence* condition as a reference. An interesting pattern emerged: all the significantly different comparisons showed advantage for conditions that only required to hold a single sentence in working memory (the *1-sentence* and *2-sentences+pre-prompt* conditions) over those that required holding two sentences in working memory (the *2-sentences-repeat both* and the *2-sentences+post prompt* conditions). More specifically, for the CI group, *2-sentences+post prompt s*cores were not only significantly lower than *1-sentence* scores (p<0.001) as indicated above, but also significantly lower than *2-sentences+pre-prompt* scores (p<0.001). The same thing happened for *2-sentence* scores, which were lower than *1-sentence* scores (p<0.01) and *2-sentences+pre-prompt* scores (p<0.05). In the normal hearing group, *2-sentences+post prompt* scores were significantly lower than the *1-sentence* scores (p<0.01) and the *2-sentences+pre-prompt scores* (p<0.05). Again, there were no significant differences between the two conditions that required holding a single sentence in memory for either the CI group or the normal hearing group, and neither were there any significant differences between the two conditions that required holding two sentences in memory, either for the CI group or the normal hearing group.,

The 2-sentence decrement, again defined as the difference in percent words correct between the *1-sentence* condition and the *2-sentences+post prompt* condition, showed large individual differences in both groups, and these were particularly pronounced in the CI group. The mean 2-sentence decrement was 13.5 percentage points for the CI group (range: −3 to 31, see Table 5) and 8 percentage points for the normal hearing group (range: 0 to 16).

**Table 5.**
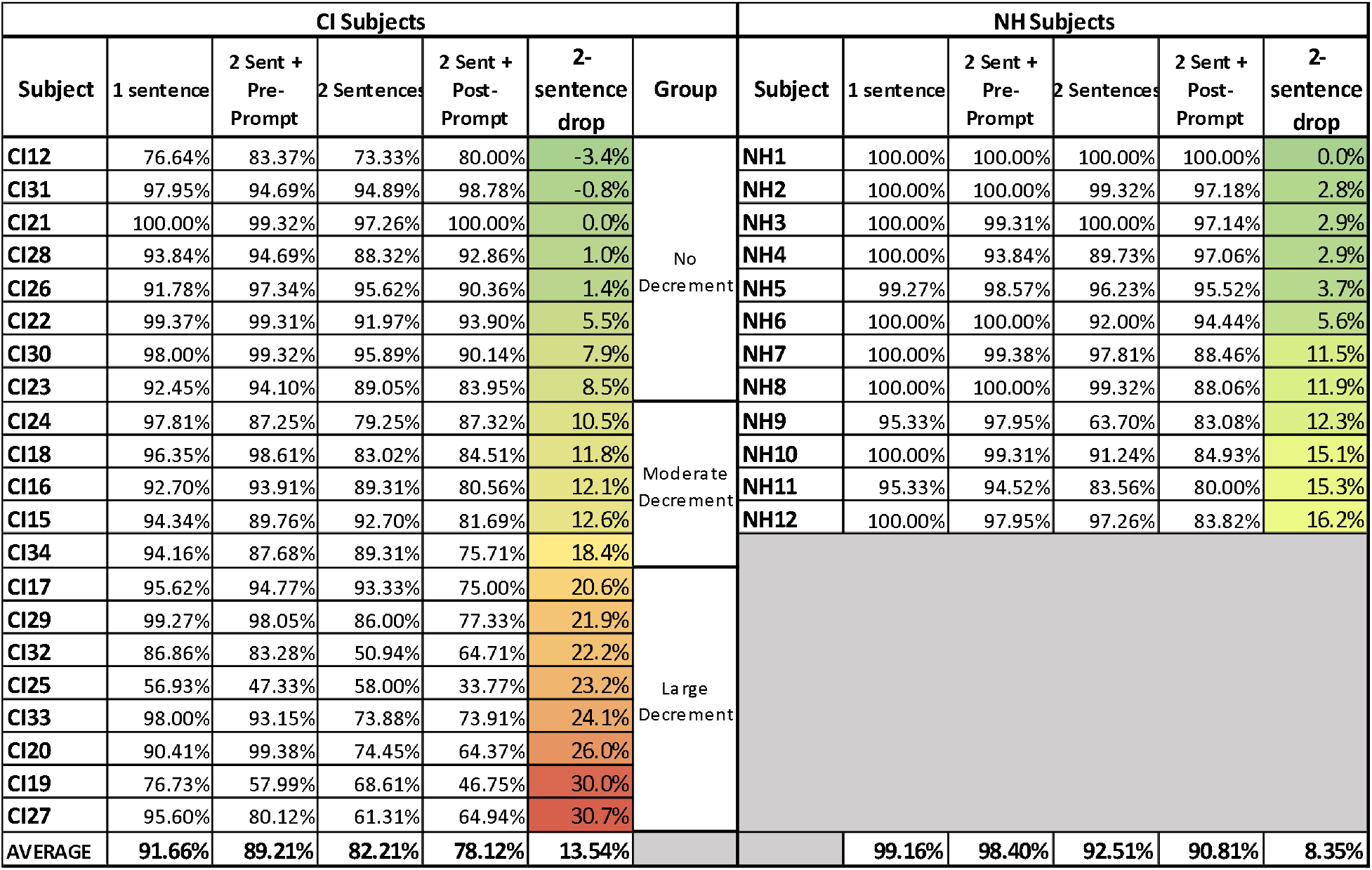
Speech scores and 2-sentence decrement, Experiment 2. Two-sentence decrements shown are based on the difference between the *1-sentence* score and the *2-sentences+post prompt* score just like Table 3 from Experiment 1. The addition of normal hearing subjects are shown as well (right side).

The variability in the 2-sentence decrement in the CI users motivated an additional analysis of their speech data. Results were analyzed for participants whose 2-sentence decrement was less than ten percentage points (the “no decrement” subgroup, N=8), between ten and twenty percentage points (the “moderate decrement” subgroup, N=5) or over 20 percentage points (the “large decrement” subgroup, N=8). Unlike previous analyses where the “group” factor referred to the CI group vs. the normal hearing group, in the present analysis all subjects were CI users and a “subgroup” factor refers to no decrement vs. moderate decrement vs. large decrement. The *2-sentences+post prompt* was not included in the analysis because it was used to determine decrement subgroup categories. These data are shown in Table 5.

A mixed-effects two-way ANOVA showed a significant main effect of decrement subgroup (F [2, 18] = 3.56, p<0.05, η_p_^2^ = 0.74) and condition (F [1.902, 34.24] = 7.514, p<0.01, η_p_^2^ = 0.56), but no significant decrement subgroup by condition interaction. For the no decrement and the moderate decrement subgroups there were no significant differences between the *1-sentence* condition and any of the other conditions, but for the large decrement subgroup there was a significant decrement of 16.6 percentage points from the *1-sentence* condition to the *2-sentences-repeat both* condition (p<0.05). This might seem like a circular result as one might expect to see a large decrement from the *1-sentence* condition to the *2-sentences-repeat both* condition in the large decrement subgroup. It is the case, however, that the large decrement subgroup was defined based on the difference from the *1-sentence* condition to the *2-sentences+post prompt* condition, while what is being discussed here is the decrement experienced in the *2-sentences-repeat both* condition. Thus, this represents a validity check indicating that the 2-sentence decrement is (at least to some extent) reproducible at the individual level. The *2-sentences-repeat both* and the *2-sentences+post prompt* scores were positively correlated (r = 0.79, p<0.001). This is also consistent with the hypothesis that there are meaningful, reproducible differences in the way individuals are affected by this 2-sentence phenomenon.

The 2-sentence decrement scores did not correlate with scores in traditional word or sentence tests (CNC30: r =-0.08, p=0.74; AzBio: r =-0.03, p=0.89), participant age (r =-0.15, p=0.52), gender (r = −0.20, p=0.39) time since first implant (r = 0.07, p=0.77), or time since the most recent implant (r =-0.04, p=0.87). Neither was the 2-sentence decrement significantly correlated with any of the variables in the baseline battery, even without correcting for multiple comparisons. As might be expected, however, scores in speech perception tests were highly correlated with each other (CNC30 vs. AzBio scores, (r = 0.91, p<0.001); CNC scores vs. the *1-sentence* condition, r = 0.67, p<0.001; AzBio scores vs. the *1-sentence* condition, r = 0.70, p<0.001); *2-sentences-repeat both* vs. *2-sentences+post prompt*, r = 0.79, p<0.001).

In summary, subjects who performed better in one speech perception test tended to perform better in other speech perception tests but no evidence was found (at least in the present data set) that 2-sentence decrement could be predicted from the scores in any of those tests, from the demographic factors we examined, or from any score from in the baseline battery.

#### Pupillometry: analysis of peak pupil dilation

Figure 3 shows the time course of pupil dilation for CI users (left panel) and normal hearing participants (right panel) for all experimental conditions, including standard error “ribbons” and behavioral percent correct scores associated with each condition.

**Figure 3.**
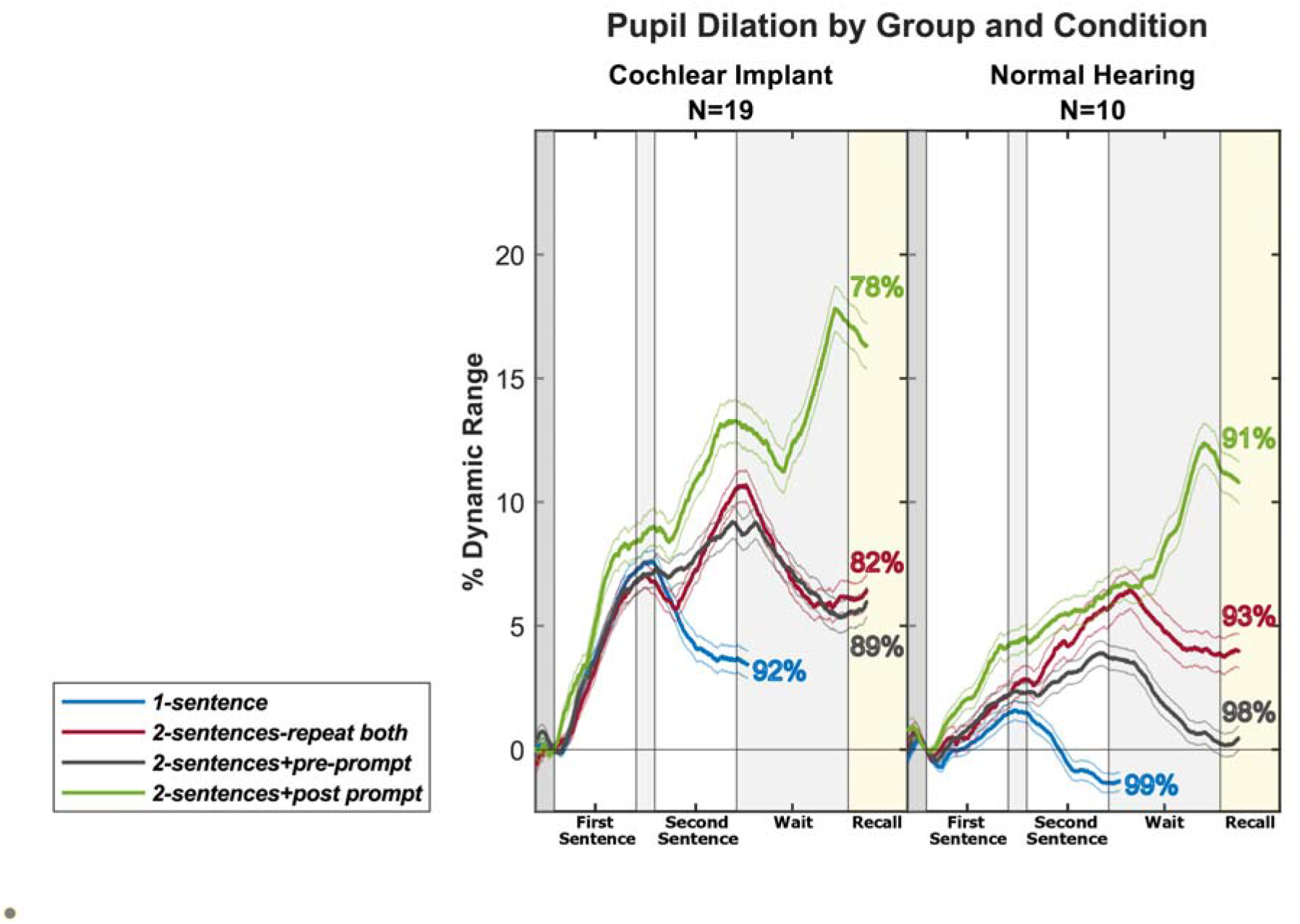
Pupil dilation for each condition expressed as percentage of dynamic range for CI users (left panel) and normal hearing listeners (right panel). The shaded areas indicate plus/minus one standard error of the mean, and color-coded numbers indicating the speech perception scores for each condition (taken from Table 5).

A mixed-effects two-way ANOVA conducted on the peak pupil dilations showed a significant effect of participant group (*F* [1, 27] = 7.295, p<0.05, η_p_^2^ = 0.46) demonstrating that peak pupil dilation, our measure of processing effort, was greater for the CI users than for the normal hearing participants. There was also a significant main effect of condition (F (2.506, 66.82) = 52.87, p<0.001, η_p_^2^ = 0.60) but no significant group by condition interaction (F (3, 80) = 0.2231, ns, η_p_^2^ = 0.02). Šídák’s multiple comparisons test shows that the group difference was significant in the *1-sentence* and the *2-sentences+pre-prompt* conditions (*p* < 0.01 in both cases).

Despite the overall difference in pupil dilation between participant groups, the pattern of change across conditions was the same for both groups: the *1-sentence* condition resulted in the lowest peak pupil dilations, the *2-sentences+post prompt* resulted in the highest peak pupil dilations, and the peak values for the *2-sentences-repeat both* and *2-sentences+pre-prompt* conditions fell in between. For the CI users, peak pupil dilation was significantly lower in the *1-sentence* condition than in the *2-sentences-repeat both* (p<0.05) and the *2-sentences+post prompt* (p<0.001) conditions, according to Dunnett’s multiple comparison test. The normal hearing group showed a similar pattern, with significant increases in peak pupil dilation in the 2-sentence condition (*p* <0.001), the *2-sentences+pre-prompt* condition (*p* <0.01), and the *2-sentences+post prompt* condition, p <0.001) with respect to the *1-sentence* condition.

A modest correlation (*r* = 0.54, p = 0.017) was found in the CI group between the 2-sentence decrement and the difference in peak pupil dilation between those two conditions. This suggests that participants who suffered greater 2-sentence decrements in speech scores when adding a second sentence to the test also showed greater increases in pupil dilation.

A secondary post-hoc analysis comparing all conditions (rather than using the *1-sentence* condition as a reference) for the CI group showed that pupil dilation in the *2-sentences+post prompt* condition was significantly higher than in all other conditions (*p*<0.001 in all cases). The same pattern was observed for the normal hearing group, with peak pupil dilation larger in the *2-sentences+post prompt* condition than in the *1-sentence* condition (*p* <0.01), the *2-sentences-repeat both* condition (p<0.05), *and the 2-sentences+pre-prompt* condition (p<0.001).

Figure 4 follows the same format as Figure 3, except that the left, middle, and right panels show three 2-sentence decrement subgroups of CI users as previously described.

**Figure 4.**
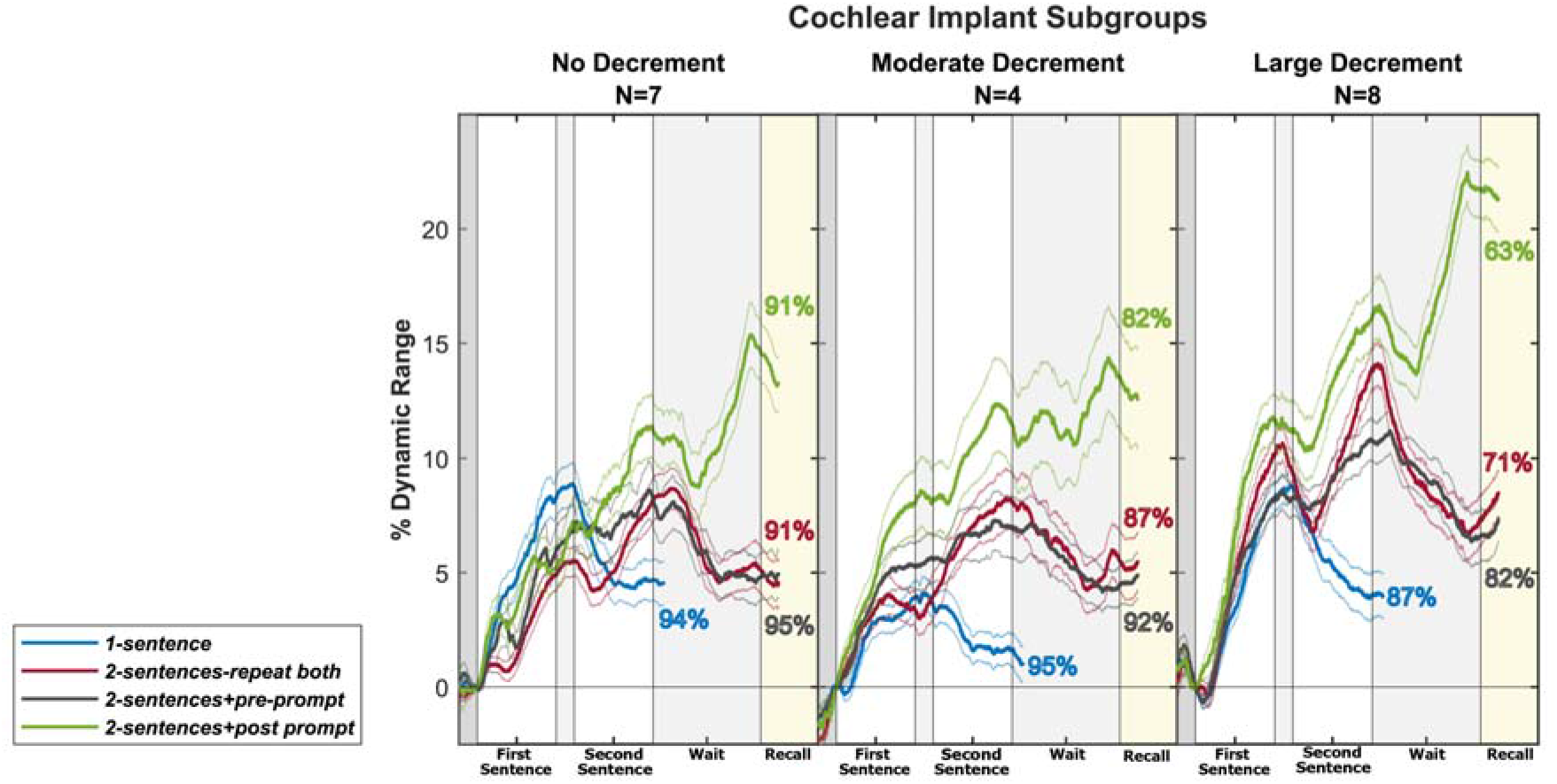
Pupil dilation for each condition expressed as percentage of dynamic range as a function of time, for the no-decrement CI subgroup (left panel), the moderate-decrement CI subgroup (middle panel), and the large-decrement CI subgroup (right panel). All other aspects of the figure are the same as in Figure 3.

These data were submitted to a mixed effects two-way ANOVA to examine the effect of condition (within-participants factor) and 2-sentence decrement subgroup (between-participants factor). The ANOVA confirmed a significant main effect of condition (F (2.379, 37.27) = 34.35, *p*< 0.001, η_p_^2^ = 0.59), and a significant condition by subgroup interaction (F (6, 47) = 3.196, p<0.05, η_p_^2^ = 0.22), but no significant effect of 2-sentence decrement subgroup. Post-hoc Dunnett’s multiple comparisons showed a significant increase in peak pupil dilation in the *2-sentences+post prompt* condition with respect to the *1-sentence* condition for the no decrement subgroup (*p* < 0.05) and for the large decrement subgroup (p< 0.001). The difference for the moderate decrement subgroup was not significant, but this subgroup had few participants.

#### Pupillometry: qualitative observations

All the preceding analyses of pupillometry data were based on peak values but it is also worth examining the time course of the different pupillometry curves. For example, Figures 3 and 4 show that the timing of the curves for each condition is perfectly consistent with the timing of the required listening effort, for both the cochlear implant and the normal hearing groups and for each cochlear implant subgroup. Peaks are typically observed right after the end of the single sentence in the *1-sentence* condition (blue curves), after the end of the second sentence in the *2-sentences-repeat both* condition (red curves) as well as in the *2-sentences+pre-prompt* condition (purple curves), and closely around the time of the prompt indicating which sentence has to be repeated in the *2-sentences+post prompt* condition (green curves). This last observation indicates that there was a clear dissociation during the wait period following sentence presentation between the *2-sentences+post prompt* condition and all the other conditions, a result that is observed for all subject groups and subgroups. Pupil dilation increases during the wait period for the *2-sentences+post prompt* condition while it decreases for all other conditions. This likely reflects the additional effort of waiting for the prompt indicating whether the first or the second sentence is to be repeated, on top of the effort already required for remembering the two sentences. The latter, but not the former, is also required in the *2-sentences-repeat both* condition, which shows a decrease in pupil dilation during the wait period.

Another interesting observation arises from comparing subsets of responses in the *2-sentences+pre-prompt* condition. In this condition, subjects are told in advance that they will have to repeat the first sentence or the second sentence. Figure 5 shows pupil dilation curves when subjects know they will only have to repeat the first sentence (purple curves) and when they know they will only have to repeat the second sentence (yellow curves). About halfway through the first sentence, subjects start showing larger pupil dilations when they are told they have to repeat the first sentence than when they are told they have to repeat the second sentence. This may reflect the increased effort associated with processing the sentence that is being presented when listeners know they will have to recall it. The fact that this difference in effort between the two conditions only becomes apparent approximately 1 second after the beginning of the sentence is consistent with the physiological lag between increased cognitive effort and pupil dilation.

**Figure 5.**
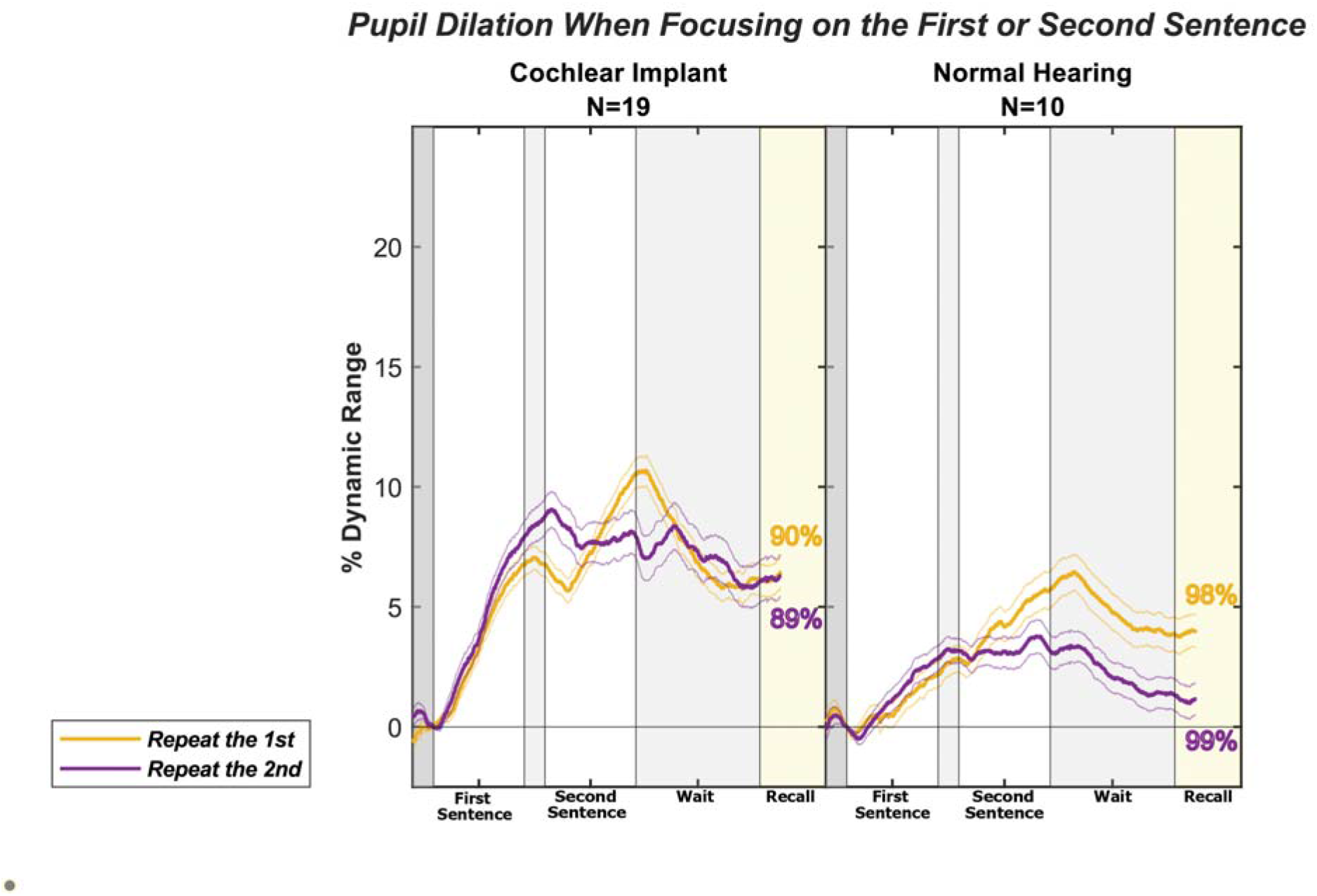
Pupil dilation expressed as a percentage of dynamic range as a function of time for the 2 sentences+pre-prompt condition. Purple and yellow curves show responses when listeners are asked (prior to stimulus presentation) to repeat only the first or the second sentence, respectively. Cochlear implant users are shown on the left panel, normal hearing listeners on the right panel. All other aspects of the figure are the same as in Figure 3.

This phenomenon is reversed, as expected, during the course of the second sentence. The purple curves remain at the same level or even start going down, possibly reflecting the decreased effort that results from disregarding auditory input after the end of the first sentence. Conversely, the yellow curves all show substantial increases during the second sentence, which are consistent with the effort required to perceive, process, and remember this sentence. These differences in pupil dilation depending on prior instructions (repeat the first or the second sentence) were consistent for both groups (CI, left panel of Figure 5, and NH, right panel, as well as for all CI subgroups, not shown).

Taken together, the preceding observations suggest that temporal aspects of the pupil dilation curves in response to different perceptual and cognitive demands represent a promising area for future studies.

#### Comparison to preregistered analyses

The preregistration document hypothesized that speech perception accuracy would be lower, and pupil dilation would be higher in the *two-sentence* and *2-sentences+post prompt* conditions than in the *1-sentence* condition. Because the preregistration was created October 18, 2018, while Experiment 1 was well underway, only data from Experiment 2 will be examined here. There are differences between the preregistration document and the actual study. Some of those are benign. What’s referred to as “Experiment 1” in the preregistration is actually a subset of Experiment 2 in the present study (the preregistration did not include the normal hearing control group or the *2-sentences+pre-prompt condition*). Likewise, the preregistration included experiments that have not been conducted yet, such as testing normal hearing listeners with vocoder-processed stimuli or manipulating the complexity of the stimulus sentences. More importantly, there are two meaningful differences between the preregistration and the present study. First, the preregistration said that a MOCA score of at least 26 would be an inclusion criterion whereas this study includes two subjects with scores of 24 and 25, respectively. Second, the preregistration indicated that 24 younger (18-45 y.o.) and 24 older (60-80 y.o.) cochlear implant users would be tested but “slow subject recruitment may require early termination of the experiment for the CI group”. This is indeed what happened, as only 21 subjects participated in Experiment 2. No formal criteria for early termination and alternative analysis paths were discussed in the preregistration so this opened the door to an important “investigator degree of freedom”, which would be to run tests for each N until a significant result was reached. This is a common practice in our field, but one that greatly increases the probability of type I errors.

To address this problem, speech perception and pupil data were re-analyzed as discussed in the preregistration after removing the two subjects with MOCA scores under 26, and then using the Pocock correction (Pocock 1977) to account for 20 multiple comparisons (for all values of N from 2 to 21). A repeated measures one-way ANOVA followed by uncorrected Fisher’s LSD test showed that speech perception scores in the *one-sentence* condition were significantly higher than those in the *two-sentence* and *2-sentences+post-prompt* conditions (p=0.0020 and p<0.0001 respectively), and results remained significant after Pocock correction (p<0.05 and p<0.01 respectively). A similar result was found for the peak pupil dilation values, although in this case data were analyzed by fitting a mixed model rather than one-way ANOVA because one of the subjects had missing data in one condition. Peak pupil dilation was significantly lower in the *1-sentence* condition than in the *2-sentences-repeat both* and *2-sentences+post prompt* conditions (p=0.0073 and p<0.0001 respectively), and results remained significant after Pocock correction (p<0.05 and p<0.01 respectively).

Thus, the hypothesis that speech perception accuracy would be lower, and pupil dilation would be higher in the *2-sentences-repeat both* and *2-sentences+post prompt* conditions than in the *1-sentence* condition has been validated by confirmatory testing. Other results in the present study should be considered exploratory.

## DISCUSSION

The main behavioral result from this study is that a simple modification of a clinical speech perception test (using two sentences at a time instead of one, to measure the “two-sentence decrement”) results in significant decreases in performance, both in normal hearing listeners and in cochlear implant users. However, the practical implications of this result are quite different for the two groups. The two-sentence decrement for normal hearing listeners is modest, and it still allows those listeners to score above 90% correct on average in all conditions. In contrast, the two-sentence decrement for the CI group averaged approximately 15 percentage points in Experiment 1 and 13.5 percentage points in Experiment 2. The practical significance of this difference is increased by the fact that CI users have a lower starting point for their *1-sentence* scores than the normal hearing listeners, and by the large individual differences within the CI group. For some CI users, the two-sentence decrement is negligible (as it is for normal hearing listeners), while for others the decrement produced by adding just a second sentence indicates potential for major difficulties in everyday communication.

The pupil dilation data tracked very closely with a commonsense assessment of required listening effort. The *1-sentence* condition requires the least effort both in terms of memory and executive function. The task in this case is simply to remember a single sentence. At the other end of the effort dimension, the *2-sentences+post prompt* condition arguably requires the most effort: the listener has to remember two sentences instead of one, wait for the prompt indicating whether the first or second sentence has to be repeated, and then provide their response. This additional effort was reflected in the larger peak pupil dilations. The other conditions are intermediate in the sense that they require greater effort than the *1-sentence* condition either in terms of memory (the *2-sentences-repeat both* condition requires remembering two sentences instead of one) or executive function (the *2-sentences+pre-prompt* requires remembering only one sentence, with a-priori knowledge that it will be the first or second one, but it also requires inhibiting processing in response to the irrelevant sentence), and they also require less effort than the *2-sentences+post prompt* condition The fact that pupil dilation tracked closely with effort at the group level is particularly interesting in the normal hearing group, which had speech perception scores near ceiling levels for all conditions.

In the introduction we mentioned the concept of a tipping point beyond which real-world oral communication becomes too difficult. There is no question that all listeners have such a tipping point, but its ecological relevance is different for people with normal hearing and those with hearing impairment. For normal hearing listeners it takes a large amount of background noise to bring them beyond a communicative tipping point, and such excessively noisy conditions are relatively rare in everyday live for most people. In contrast, adult CI users, even those who are deemed “good performers” in clinical tests of speech perception, may be brought beyond the tipping point by a wide range of ecologically relevant manipulations. In this study we document that this happens to many (but not all) CI users even when speech input is only minimally more challenging than standard clinical tests: using two sentences instead of a single sentence or a single word.

The present findings raise the possibility that a clinical version of a 2-sentence test may provide actionable information for counseling and rehabilitating CI users. For example, a patient who does very well with standard clinical speech tests may be less likely to be referred for aural rehabilitation under the current standard of care, but a sizeable 2-sentence decrement may persuade their clinician to recommend such rehabilitation nonetheless. It is likely that the interaction of limited cognitive resources and a highly degraded auditory input manifests itself in many other situations for this important clinical population, and the study of this interaction represents a fruitful area for future research.

## Data Availability

All data produced in the present study are available upon reasonable request to the authors.

## ACKNOWLEDGMENTS

This work was supported by the National Institutes of Health (NIH) Grant No. R01 DC016834 from the National Institute of Deafness and Other Communication Disorders. We are grateful to Alexander Kinney for his help setting up the EyeLink device at the NYU site. We also gratefully acknowledge support from the Stephen J. Cloobeck Research Fund.

Preregistered on AsPredicted #15259

This manuscript is available in preprint at MedRxiv. MEDRXIV/2022/277720

